# Metformin and Severe Post-COVID-19 Outcomes Among Individuals with Diabetes Mellitus

**DOI:** 10.64898/2026.07.06.26357398

**Authors:** Zachary Butzin-Dozier, Lin-Chiun Wang, Yunwen Ji, A. Jerrod Anzalone, Oluwasolape Olawore, Ryan Hafen, Eric Hurwitz, Manav Kumar, Rena C. Patel, Ariana Budhihartanto, Mark van der Laan, John M. Colford, Alan E. Hubbard, John B. Buse, Steven Johnson, Jane Reusch, Lauren E. Chan, Richard Moffitt, Rachel Wong, Carolyn Bramante, the National Clinical Cohort Collaborative (N3C) Consortium

## Abstract

**Background:** Metformin is one of the most commonly prescribed medications for individuals with diabetes and may provide protection against long-term sequelae of COVID-19.

**Methods:** We evaluated a retrospective cohort of individuals in the National Clinical Cohort Collaborative with type 2 diabetes mellitus and COVID-19 who were prescribed metformin or a dipeptidyl peptidase-4 inhibitor (DPP4i) at least 30 days before the onset of acute COVID-19 between October 1, 2021, and November 15, 2023. We compared the 12-month cumulative incidence of Long COVID diagnosis (ICD-10 U09.9: Post COVID-19 condition, unspecified), probable Long COVID (based on a model-derived phenotype), and mortality between individuals prescribed metformin vs. DPP4i. We applied Super Learner and targeted maximum likelihood estimation to obtain risk ratios while adjusting for covariates of interest.

**Results:** In our sample of 53,332 individuals with type 2 diabetes and COVID-19, we found that metformin prescription was associated with a lower risk of all-cause mortality after COVID-19 (adjusted risk ratio [aRR] 0.61, 95% CI 0.51, 0.73). We also observed that metformin users, compared to DPP4i users, had a slightly lower risk of probable Long COVID (aRR 0.87, 95% CI 0.81, 0.94) but did not detect a significant relationship with Long COVID diagnosis (aRR 0.90, 95% CI 0.68, 1.20), although we observed similar point estimates across Long COVID outcomes.

**Conclusions:** These findings support the hypothesis that metformin prescription during acute COVID-19 may be associated with lower mortality among adults with diabetes. These analyses also provide modest evidence of a protective association against Long COVID in adults with diabetes, although estimates were imprecise.

---

Long COVID, also known as post-acute sequelae of COVID-19 or long-haul COVID, is a broad array of conditions and symptoms that individuals develop after the initial SARS-CoV-2 infection subsides.^1–3^ Individuals with diabetes are at increased risk of mortality after respiratory virus infections, and of post-infectious syndromes like Long COVID.^4–6^ Type 2 diabetes mellitus (T2DM) results from a combination of ineffective insulin secretion by pancreatic β-cells and reduced insulin sensitivity, and it is one of the most common metabolic disorders worldwide.^7,8^ Metformin, a first-line treatment for T2DM, is an antihyperglycemic medication with considerable immunomodulatory effects and host-mediated antiviral properties that may protect individuals against the long-term sequelae of COVID-19.^9–13^ Evaluating the relationship between antihyperglycemic medication use and severe post-COVID-19 outcomes may inform clinical management strategies for individuals with T2DM, a population at elevated risk for severe COVID-19 complications.

Randomized controlled trials have evaluated the impact of metformin on COVID-19 severity and the incidence of Long COVID.^9,14,15^ The COVID-OUT trial found that outpatient metformin use during acute SARS-CoV-2 infection was associated with lower odds (odds ratio 0.58, 95% confidence interval (CI) 0.35 to 0.94) of emergency department visit, hospitalization, or death, and 0.59 times the hazard (95% CI 0.39 to 0.89) of Long COVID.^11,15^ The ACTIV-6 randomized controlled trial found modest evidence that assignment to metformin, versus placebo, was associated with a decreased risk of Long COVID diagnosis over 6 months (adjusted risk ratio 0.50, 95% credible interval 0.16 to 1.00), but was not associated with increased Long COVID symptoms (adjusted risk ratio 0.79, 95% credible interval 0.47 to 1.23).^10^ Observational studies of individuals in the National Clinical Cohort Collaborative (N3C) have provided additional insights. Metformin was associated with a lower risk of mortality, hospitalization, and mechanical ventilation in individuals with T2DM and acute COVID-19.^16^ In a cohort of individuals with prediabetes, investigators found that metformin use, compared with levothyroxine or ondansetron, was associated with decreased acute COVID-19 severity as well as decreased mortality.^9^ In a cohort of individuals with T2DM, investigators found that metformin use, compared to sulfonylureas (SU) use, at the time of acute COVID-19 was associated with 0.56 times the risk of mortality (95% CI 0.33 to 0.97).^14^ Cumulatively, these results support the hypothesis that metformin use may reduce severe sequelae of COVID-19, such as Long COVID and mortality, although additional research is needed.

While previous studies have provided considerable evidence regarding the impact of metformin on acute COVID-19 and Long COVID, several questions remain. Randomized trials often suffer from poor generalizability, as trial populations often differ from the general population. In a cohort of individuals with T2DM in N3C, we aim to evaluate whether the prescription of metformin is associated with a lower risk of Long COVID and mortality, compared with the prescription of dipeptidyl peptidase-4 inhibitors (DPP4i). In two secondary cohorts (see Supplemental Material 1 for rationale), we evaluated the relationships between metformin, Long COVID, and mortality in individuals with (1) PCOS and (2) prediabetes to evaluate the generalizability of our findings in individuals with insulin resistance.

## METHODS

### Data source

We analyzed data from the N3C, the largest open, national source of health data in U.S. history, containing data on 21 million individuals from 82 data providers.^17,18^ N3C includes electronic medical record data from participants ranging from January 2018 to the present.

### Sample

We included individuals in N3C with type 2 diabetes mellitus (T2DM) aged 30-85 years who were diagnosed with COVID-19 between October 1, 2021 (the release date of the ICD code U09.9, Post COVID-19 condition, unspecified) and November 15, 2023, and were prescribed either metformin or DPP4i during (prevalent use) acute COVID-19 (metformin and DPP4i will hereafter be referred to as “study drugs”).^14^ We excluded individuals who (A) were first prescribed a study drug less than 30 days before acute COVID-19, (B) were prescribed multiple study drugs within 30 days before acute COVID-19, or (C) had a prescription end date before acute COVID-19. We did not exclude patients based on medication history more than 30 days before acute COVID-19, and we did not incorporate treatment change after acute COVID-19, following an emulated intention-to-treat design. The T2DM cohort included individuals with either a diagnosis code for T2DM or a hemoglobin A1C (HbA1C) greater than 6.5%. We excluded individuals diagnosed with any of the following conditions at baseline: chronic kidney disease stages 3, 4, or 5; end-stage renal disease; prediabetes (analyzed separately as a secondary prediabetes cohort); or polycystic ovarian syndrome (analyzed separately as a secondary PCOS cohort).^14^ We excluded individuals who were prescribed one of the study drugs as treatment for severe acute COVID-19 to avoid confounding by indication.

### Exposure

The exposure of interest was the prescription of metformin compared to DPP4i, which is an alternative or adjunct pharmacotherapy to metformin and which has been demonstrated to have some reduction in COVID-19 severity.^14^

### Outcomes

The outcomes of interest were the cumulative incidence of a Long COVID diagnosis (LC-Dx (defined by ICD-10 code U09.9: Post COVID-19 condition, unspecified), probable Long COVID (LC-P), or mortality within 12 months following the prescription of diabetes treatment medication. The outcome of interest, Long COVID, was defined using 2 different methods: 1) LC-Dx: diagnosis code U09.9, which became available in 10/2021 (LC-Dx), and to address limited capture of the Long COVID due to the poor early update of diagnosis code use 2) LC-P: a validated model-based phenotype based on patient conditions, symptoms, diagnoses, and laboratory measures.^19–23^ We considered a patient as having “probable Long COVID” if the patient had a Long COVID model-based phenotype score at or above 0.9 in 1-12 months following acute COVID-19. Previous studies have used 0.9 as a threshold for LC-P using the N3C Long COVID model-based phenotype.^24,25^

### Confounders and other covariates

We included the following individual baseline covariates in our analyses based on potential relationships with the exposures and outcomes of interest (where the baseline is prior to prescription of study drug): healthcare utilization rate (healthcare interactions per month), sex, age, race and ethnicity, data provider, body mass index, tobacco use, medical conditions (obesity, chronic lung disease, hypertension, asthma, heart failure, dementia, arthritis, coronary artery disease, cancer, liver disease, chronic kidney disease, peripheral vascular disease, cerebrovascular disease, polycystic ovarian syndrome (PCOS), and depression), medication use (systemic corticosteroids, outpatient insulin, angiotensin converting enzyme (ACE) inhibitors, angiotensin receptor blockers, statins, anticoagulants, aspirin, torsemide, and furosemide), biomarker measurements (glycated hemoglobin A1c (A1c), serum creatinine, urine albumin to creatinine ratio, and estimated glomerular filtration rate [reported by EHR laboratory measurements]), and we tracked patient monitoring (frequency of healthcare interactions during baseline and follow-up period) (detailed covariate definitions published previously,^26^ covariate list included in Supplemental Material 2). For medical conditions and medication use, we evaluated any history between January 2018 (beginning of N3C observation period) and acute COVID-19. For biomarker measurements and BMI, we included the most recent value before acute COVID-19. We intervened on monitoring to evaluate the counterfactual impact of the treatment of interest, given that all individuals are monitored (healthcare interaction) once every six months during the study period.

### Secondary cohort

As a secondary research question, we evaluated the relationship between metformin use, compared to levothyroxine, and Long COVID or mortality in cohorts of individuals with (A) prediabetes and (B) PCOS (excluding individuals with both prediabetes and PCOS). We selected levothyroxine as a comparator for our secondary cohort as it is a commonly prescribed oral medication (i.e., requires healthcare utilization) with no known antihyperglycemic or protective effects against COVID-19.^9,14^

### Analysis

Our analysis applied Super Learner to maximize the prediction of the outcomes of interest, Long COVID and mortality, given individual treatment and covariate status, as well as the treatment mechanism (probability of prescription to metformin, given covariate information) and censoring mechanism (probability of loss to follow-up, given covariate and treatment information).^27–29^ Super Learner creates a convex combination of candidate algorithms and is guaranteed to perform at least as well as the best-fitting candidate algorithm in large sample sizes. Super Learner is ideal for this data setting, as high-dimensional covariate information will inevitably lead to model misspecification in traditional parametric analyses. Next, we used targeted maximum likelihood estimation to reduce bias and estimate the risk ratio comparing metformin use with DPP4i use for the one-year cumulative incidence of LC-Dx, LC-P, and mortality.^30–34^ Targeted maximum likelihood estimation is a doubly robust method that yields consistent inferences as long as the outcome regression or the treatment mechanism is estimated consistently. In this data setting, where the treatment decisions regarding metformin vs. DPP4i are difficult to characterize, this approach provides additional robustness against near positivity violations compared with inverse probability of treatment weighting alone.^30–32,34^ We considered the observation (healthcare interaction) as informative censoring (i.e., the censoring mechanism), estimating the counterfactual impact of the exposure (metformin prescription) under a scenario of universal observation.^27^ This counterfactual intervention included ensuring at least one healthcare interaction during the 12-month follow-up period, and for Long COVID outcomes, no mortality during the follow-up period.

## RESULTS

We evaluated EHR data from 53,332 individuals with T2DM, including 50,965 individuals who were prescribed metformin during acute COVID-19 and 2,367 who were prescribed DPP4i during acute COVID-19 (Table 1). In our sample, 52% of metformin users were female, while 59% of DPP4i users were female. The average age of metformin users was 60 years, while the average age of DPP4i users was 64 years. The mean BMI for metformin users was 36, and for DPP4i users, 35. The average baseline HbA1c was 7.7% among metformin users and 7.9% among DPP4i users. Metformin users had an average of 2.6 interactions with healthcare providers per month, whereas DPP4i users had an average of 2.7 interactions per month.

**Table 1.** Characteristics of individuals with type 2 diabetes prescribed metformin or DPP4i during acute COVID-19.

| Characteristic | Value | Metformin Count<br>(Proportion) | DPP4i Count<br>(Proportion) | Total Count<br>(Proportion) |
| --- | --- | --- | --- | --- |
| Total |  | 50965 (0.96) | 2367 (0.04) | 53332 (1) |
| Sex | Female | 26694 (0.52) | 1389 (0.59) | 28083 (0.53) |
| Age: mean (SD) |  | 60.23 (12.16) | 63.51 (11.96) | 60.37 (12.17) |
| Ethnicity | White Non-Hispanic | 29353 (0.58) | 1314 (0.56) | 30667 (0.58) |
|  | Black or African American Non-Hispanic | 8853 (0.17) | 491 (0.21) | 9344 (0.18) |
|  | Hispanic or Latino Any Race | 6761 (0.13) | 311 (0.13) | 7072 (0.13) |
|  | Unknown | 2043 (0.04) | 90 (0.04) | 2133 (0.04) |
|  | Asian or Pacific Islander Non-Hispanic | 3372 (0.07) | 137 (0.06) | 3509 (0.07) |
| BMI: mean (SD) |  | 36.32 (8.68) | 34.91 (8.25) | 36.26 (8.67) |
| Medical Conditions | Tobacco Smoker | 8785 (0.17) | 399 (0.17) | 9184 (0.17) |
|  | Obese | 36375 (0.71) | 1545 (0.65) | 37920 (0.71) |
|  | Chronic Lung Disease | 12683 (0.25) | 685 (0.29) | 13368 (0.25) |
|  | Hypertension | 38033 (0.75) | 1737 (0.73) | 39770 (0.75) |
|  | Systemic Corticosteroids | 23354 (0.46) | 1095 (0.46) | 24449 (0.46) |
|  | Asthma | 8399 (0.16) | 395 (0.17) | 8794 (0.16) |
|  | Heart Failure | 5286 (0.1) | 308 (0.13) | 5594 (0.1) |
|  | Dementia | 1042 (0.02) | 62 (0.03) | 1104 (0.02) |
|  | Arthritis | 1166 (0.02) | 66 (0.03) | 1232 (0.02) |
|  | Coronary Artery Disease | 8806 (0.17) | 476 (0.2) | 9282 (0.17) |
|  | Cancer | 6404 (0.13) | 332 (0.14) | 6736 (0.13) |
|  | Liver Disease | 779 (0.02) | 58 (0.02) | 837 (0.02) |
|  | Chronic Kidney Disease | 4573 (0.09) | 353 (0.15) | 4926 (0.09) |
|  | Peripheral Vascular Disease | 6809 (0.13) | 372 (0.16) | 7181 (0.13) |
|  | Cerebrovascular Disease | 3812 (0.07) | 227 (0.1) | 4039 (0.08) |
|  | Depression | 13827 (0.27) | 642 (0.27) | 14469 (0.27) |
| Drugs | Insulin | 21255 (0.42) | 1214 (0.51) | 22469 (0.42) |
|  | ACE Inhibitors | 538 (0.01) | 33 (0.01) | 571 (0.01) |
|  | Statins | 1010 (0.02) | 58 (0.02) | 1068 (0.02) |
|  | Anticoagulants | 2084 (0.04) | 110 (0.05) | 2194 (0.04) |
|  | Aspirin | 1548 (0.03) | 83 (0.04) | 1631 (0.03) |
|  | Torsemide | 486 (0.01) | 35 (0.01) | 521 (0.01) |
|  | Furosemide | 6147 (0.12) | 392 (0.17) | 6539 (0.12) |
| Measurements | Pre-Prescription HbA1c: mean (SD) | 7.65 (1.81) | 7.93 (1.88) | 7.66 (1.81) |
|  | Serum Creatinine: mean (SD) | 0.87 (0.25) | 0.95 (0.4) | 0.88 (0.26) |
|  | Albumin/Creatinine Ratio: mean (SD) | 10.84 (36.45) | 13.21 (39.74) | 10.95 (36.61) |
|  | Estimated Glomerular Filtration Rate: mean (SD) | 77.19 (21.12) | 71.27 (23.08) | 76.92 (21.25) |
| Medical Utilization | Visits per Month: mean (SD) | 2.63 (3.06) | 2.68 (3.39) | 2.63 (3.07) |

We found that prescription of metformin, compared with DPP4i, during acute COVID-19 was associated with lower 12-month mortality risk (adjusted risk ratio (aRR) 0.61, 95% CI 0.51, 0.73; unadjusted risk ratio (uRR) 0.44, 95% CI 0.37, 0.53) and 12-month risk of LC-P (aRR 0.87, 95% CI 0.81, 0.94; uRR 0.85, 95% CI 0.78, 0.92) (Figure 1, Table 2). We did not find that prescription of metformin, compared to DPP4i, was significantly associated with 12-month cumulative incidence of Long COVID diagnosis (aRR 0.90, 95% CI 0.68, 1.20; uRR 0.80, 95% CI 0.57, 1.11)).

**Figure 1.**
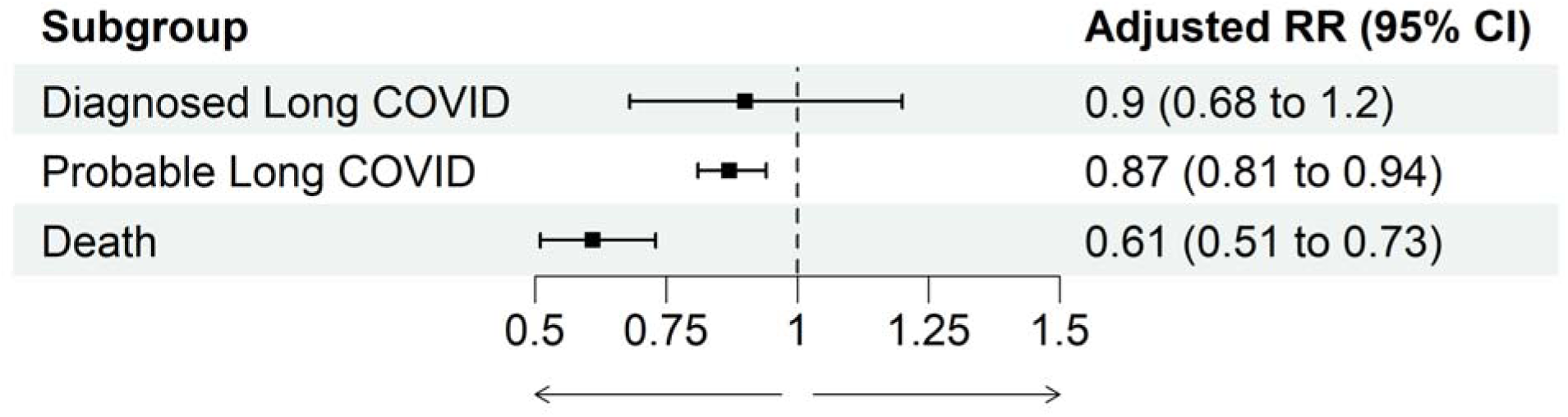
Adjusted relationships between metformin vs. DPP4i during acute COVID-19 and subsequent 12-month cumulative incidence of Long COVID or death, among individuals with type 2 diabetes mellitus.

**Table 2.** Unadjusted relationships between metformin vs. DPP4i during acute COVID-19 and subsequent 12-month cumulative incidence of Long COVID or death, among individuals with type 2 diabetes mellitus.

| Outcome | Metformin Sample Size | DPP4i Sample Size | Count Metformin Outcomes | Count DPP4i Outcomes | Metformin Unadjusted Risk | DPP4i Unadjusted Risk | Unadjusted RR (95% CI) |
| --- | --- | --- | --- | --- | --- | --- | --- |
| Diagnosed Long COVID | 50965 | 2367 | 635 | 37 | 0.01 | 0.02 | 0.80 (0.57 to 1.11) |
| Probable Long COVID | 50965 | 2367 | 8444 | 464 | 0.17 | 0.20 | 0.85 (0.78 to 0.92) |
| Death | 50965 | 2367 | 1220 | 128 | 0.02 | 0.05 | 0.44 (0.37 to 0.53) |

We evaluated EHR data from 15,582 individuals with prediabetes and 1,786 individuals with PCOS (see Supplemental Materials 3 and 4 for details on the prediabetes and PCOS cohorts). Among individuals with prediabetes or PCOS, we found that prescription of metformin, vs. levothyroxine, was not significantly associated with the one-year cumulative incidence of Long COVID diagnosis (prediabetes aRR 0.88, 95% CI 0.65 to 1.20; PCOS aRR 0.95, 95% CI 0.45 to 1.98), LC-P (prediabetes aRR 0.97, 95% CI 0.90 to 1.04; PCOS aRR 0.92, 95% CI 0.77 to 1.11), or mortality (prediabetes aRR 0.74, 95% CI 0.42 to 1.30 [PCOS invalid RR]) (see Supplemental Material 5).

## DISCUSSION

We found evidence that prescription of metformin, compared with DPP4i, was associated with lower mortality in T2DM individuals (aRR 0.61, 95% CI 0.51, 0.73). Our findings are consistent with the COVID-OUT trial, which randomized patients with COVID-19 to receive metformin, and found that metformin use during acute COVID-19 was associated with 0.58 times the odds (95% CI 0.35 to 0.94) of emergency department visit, hospitalization, or death. ^11^ Similarly, our findings were consistent with an observational study of individuals in N3C, which found that metformin use, compared to sulfonylureas, was associated with 0.56 times the risk of mortality (95% CI 0.33 to 0.97).^14^ The consistency of these findings across multiple study designs, including randomized trials and observational analyses using both prevalent and incident exposure definitions with different active comparators, supports the association between metformin use and lower mortality among individuals with T2DM and COVID-19. For discussion of secondary cohort findings, see Supplemental Material 6.

We found moderate evidence of a relationship between metformin prescription, compared to DPP4i prescription, and Long COVID among T2DM individuals. We observed similar point estimates for LC-Dx (aRR 0.90) and LC-P (aRR 0.87), although our estimate of LC-Dx had low precision due to LC-Dx being a rare outcome, with a cumulative incidence of 1% among individuals prescribed metformin and 2% among controls. This low diagnostic rate may be attributed to the wide range of Long COVID phenotypes or the lack of meaningful treatments for Long COVID, which may discourage formal diagnosis of the condition.^19,21,35^ Therefore, we included LC-P as an additional outcome, which uses a model-based phenotype and has greater sensitivity than LC-P. In contrast, LC-P may have lower clinical utility than LC-Dx, as it relies on myriad patient characteristics and conditions to approximate underlying disease status but lacks clear clinical application. The inclusion of both measures serves to evaluate the consistency of the relationships between these outcomes. Our observed point estimates were considerably more modest than previous studies, as the COVID-OUT trial found that assignment to metformin within 3 days of COVID-19 onset reduced the hazard of Long COVID (hazard ratio 0.59, 95% CI 0.39 to 0.89).^15^ The attenuated relationship observed in our study, compared to the COVID-OUT trial, may be due to residual confounding of the relationship between metformin prescription and LC-Dx and LC-P by comorbidity burden, differential monitoring, or healthcare utilization. These results add to the mixed findings of the cumulative literature, which have shown heterogeneous impacts of metformin on Long COVID depending on the definition of Long COVID used.^10,11,15^

Metformin may protect individuals with T2DM and acute COVID-19 from mortality and Long COVID through similar mechanisms. In addition to its antihyperglycemic effects, metformin has shown anti-inflammatory properties and modulates interleukin-6 and interleukin-1, which are key cytokines that trigger hyperinflammation in severe COVID-19 and Long COVID.^11,36–39^ Furthermore, metformin has demonstrated antiviral activity against SARS-CoV-2, which may improve recovery from COVID-19 and prevent Long COVID.^11,40,41^

### Strengths and Limitations

The major limitation of our study is that we did not address the effects of initiating these drug classes during the acute COVID infection, thus limiting the causal interpretation of our findings. The relatively small proportion (0.04) of T2DM individuals using DPP4i in our sample leads to imprecise estimates due to low power, particularly regarding rare outcomes like Long COVID diagnosis, which had only a 1% cumulative incidence over study period). Residual confounding by indication remains a concern, as metformin is considered a first-line treatment for T2DM patients, while DPP4i is typically a second-line or add-on therapy.^42,43^ Therefore, DPP4i patients may be less amenable to antihyperglycemic medications or may have advanced T2DM characteristics that are not captured in the EHR. The generalizability of the N3C sample is another limitation. While N3C is a large, national sample, it oversamples individuals from research institutions and individuals who frequently interact with the healthcare system, resulting in a sample population that is disproportionately high-income, white, older, and with high comorbidities.^19,21,22^

A major strength of this study was the data source, which comprised a large sample of individuals from 84 healthcare institutions across the U.S.^18^ Similarly, we included a wide range of health characteristics from each patient, allowing us to rigorously adjust for a wide range of potential confounders. The analytic approach that we applied, using Super Learner and Targeted Maximum Likelihood Estimation, was a second strength.^30–34^ The use of highly flexible, semiparametric estimation methods minimized the probability of model misspecification while conducting high-dimensional covariate adjustment, and the use of doubly-robust estimation methods supports the validity of our findings.

## CONCLUSIONS

We found that the prescription of metformin, compared to DPP4i, during acute COVID-19 was associated with lower 12-month mortality among individuals with T2DM. We found modest evidence of a protective relationship between metformin and Long COVID, although the estimation of Long COVID diagnosis was imprecise. These findings support the use of metformin in individuals with T2DM during acute COVID-19 to reduce long-term adverse outcomes.

## Data Availability

All analytic code is available upon request from the N3C Enclave. Access to study data may be requested in the N3C Enclave as legacy data pending N3C approval. Access to the N3C Data Enclave is managed by NCATS. Interested researchers must first complete a data use agreement, and next a data use request, in order to access the N3C Data Enclave. Once access is granted, the N3C data use committee must review and approve all use of data and the publication committee must approve all publications involving N3C data.

https://ncats.nih.gov/research/research-activities/n3c/resources/data-access

## SUPPLEMENTAL MATERIALS

Supplemental Material 1. Rationale for secondary analyses.

Supplemental Material 2. Covariates of interest.

Supplemental Material 3. Characteristics of individuals with prediabetes prescribed metformin or levothyroxine during acute COVID-19.

Supplemental Material 4. Characteristics of individuals with PCOS prescribed metformin or levothyroxine during acute COVID-19.

Supplemental Material 5. Relationship between metformin vs. levothyroxine during acute COVID-19 and subsequent 12-month cumulative incidence of Long COVID or death, among individuals with PCOS or prediabetes.

Supplemental Material 6. Discussion of secondary cohort findings.

S

### Ethics approval and consent to participate

This study was approved by the UC Berkeley Office for Protection of Human Subjects (2022-01-14980). The N3C data transfer to NCATS is performed under a Johns Hopkins University Reliance Protocol # IRB00249128 or individual site agreements with NIH. N3C received a waiver of consent from the NIH Institutional Review board and allows the secondary analysis of these data without additional consent.

### Consent to publish

The authors consent to the publication of this manuscript in its entirety.

Availability of data and materials: All analytic code is available upon request from the N3C Enclave. Access to study data may be requested in the N3C Enclave as “legacy data” pending N3C approval. Access to the N3C Data Enclave is managed by NCATS (https://ncats.nih.gov/research/research-activities/n3c/resources/data-access). Interested researchers must first complete a data use agreement, and next a data use request, in order to access the N3C Data Enclave. Once access is granted, the N3C data use committee must review and approve all use of data and the publication committee must approve all publications involving N3C data.

### Funding

This research was financially supported by the National Institute for Allergy and Infectious Diseases (1K01AI182501 to Zachary Butzin-Dozier). Individual authors were supported by the following funding sources: National Institute of Mental Health R01131542 (PI Rena C. Patel), Jerrod Anzalone is supported by the National Institute of General Medical Sciences, U54 GM115458, which funds the Great Plains IDeA-CTR Network. The content is solely the responsibility of the authors and does not necessarily represent the official views of the NIH.

## N3C Attribution

The analyses described in this manuscript were conducted with data or tools accessed through the NCATS N3C Data Enclave https://covid.cd2h.org and N3C Attribution & Publication Policy v 1.2-2020-08-25b supported by NCATS U24 TR002306, Axle Informatics Subcontract: NCATS-P00438-B, the Bill & Melinda Gates Foundation: OPP1165144, and the National Institutes of General Medical Sciences: U54GM115458 and 5U54GM104942-04. This research was possible because of the individuals whose information is included within the data and the organizations (https://ncats.nih.gov/n3c/resources/data-contribution/data-transfer-agreement-signatories) and scientists who have contributed to the ongoing development of this community resource [https://doi.org/10.1093/jamia/ocaa196].

## Disclaimer

The N3C Publication committee confirmed that this manuscript (MSID 2766.892) complies with N3C data use and attribution policies; however, the authors are solely responsible for its content, which does not necessarily represent the official views of the National Institutes of Health or the N3C program.

## IRB

The N3C data transfer to NCATS is performed under a Johns Hopkins University Reliance Protocol # IRB00249128 or individual site agreements with NIH. The N3C Data Enclave is managed under the authority of the NIH; information can be found at https://ncats.nih.gov/n3c/resources.

This research project was approved by the University of California, Berkeley Committee for the Protection of Human Subjects (CPHS protocol number 2022-01-14980). This approval is issued under University of California, Berkeley Federalwide Assurance #00006252.

## Individual Acknowledgements For Core Contributors

We gratefully acknowledge the following core contributors to N3C:

Adam B. Wilcox, Adam M. Lee, Alexis Graves, Alfred (Jerrod) Anzalone, Amin Manna, Amit Saha, Amy Olex, Andrea Zhou, Andrew E. Williams, Andrew Southerland, Andrew T. Girvin, Anita Walden, Anjali A. Sharathkumar, Benjamin Amor, Benjamin Bates, Brian Hendricks, Brijesh Patel, Caleb Alexander, Carolyn Bramante, Cavin Ward-Caviness, Charisse Madlock-Brown, Christine Suver, Christopher Chute, Christopher Dillon, Chunlei Wu, Clare Schmitt, Cliff Takemoto, Dan Housman, Davera Gabriel, David A. Eichmann, Diego Mazzotti, Don Brown, Eilis Boudreau, Elaine Hill, Elizabeth Zampino, Emily Carlson Marti, Emily R. Pfaff, Evan French, Farrukh M Koraishy, Federico Mariona, Fred Prior, George Sokos, Greg Martin, Harold Lehmann, Heidi Spratt, Hemalkumar Mehta, Hongfang Liu, Hythem Sidky, J.W. Awori Hayanga, Jami Pincavitch, Jaylyn Clark, Jeremy Richard Harper, Jessica Islam, Jin Ge, Joel Gagnier, Joel H. Saltz, Joel Saltz, Johanna Loomba, John Buse, Jomol Mathew, Joni L. Rutter, Julie A. McMurry, Justin Guinney, Justin Starren, Karen Crowley, Katie Rebecca Bradwell, Kellie M. Walters, Ken Wilkins, Kenneth R. Gersing, Kenrick Dwain Cato, Kimberly Murray, Kristin Kostka, Lavance Northington, Lee Allan Pyles, Leonie Misquitta, Lesley Cottrell, Lili Portilla, Mariam Deacy, Mark M. Bissell, Marshall Clark, Mary Emmett, Mary Morrison Saltz, Matvey B. Palchuk, Melissa A. Haendel, Meredith Adams, Meredith Temple-O’Connor, Michael G. Kurilla, Michele Morris, Nabeel Qureshi, Nasia Safdar, Nicole Garbarini, Noha Sharafeldin, Ofer Sadan, Patricia A. Francis, Penny Wung Burgoon, Peter Robinson, Philip R.O. Payne, Rafael Fuentes, Randeep Jawa, Rebecca Erwin-Cohen, Rena Patel, Richard A. Moffitt, Richard L. Zhu, Rishi Kamaleswaran, Robert Hurley, Robert T. Miller, Saiju Pyarajan, Sam G. Michael, Samuel Bozzette, Sandeep Mallipattu, Satyanarayana Vedula, Scott Chapman, Shawn T. O’Neil, Soko Setoguchi, Stephanie S. Hong, Steve Johnson, Tellen D. Bennett, Tiffany Callahan, Umit Topaloglu, Usman Sheikh, Valery Gordon, Vignesh Subbian, Warren A. Kibbe, Wenndy Hernandez, Will Beasley, Will Cooper, William Hillegass, Xiaohan Tanner Zhang. Details of contributions available at covid.cd2h.org/core-contributors

## Data Partners with Released Data

The following institutions whose data is released or pending:

Available: Advocate Health Care Network — UL1TR002389: The Institute for Translational Medicine (ITM) • Boston University Medical Campus — UL1TR001430: Boston University Clinical and Translational Science Institute • Brown University — U54GM115677: Advance Clinical Translational Research (Advance-CTR) • Carilion Clinic — UL1TR003015: iTHRIV Integrated Translational health Research Institute of Virginia • Charleston Area Medical Center — U54GM104942: West Virginia Clinical and Translational Science Institute (WVCTSI) • Children’s Hospital Colorado — UL1TR002535: Colorado Clinical and Translational Sciences Institute • Columbia University Irving Medical Center — UL1TR001873: Irving Institute for Clinical and Translational Research • Duke University — UL1TR002553: Duke Clinical and Translational Science Institute • George Washington Children’s Research Institute — UL1TR001876: Clinical and Translational Science Institute at Children’s National (CTSA-CN) • George Washington University — UL1TR001876: Clinical and Translational Science Institute at Children’s National (CTSA-CN) • Indiana University School of Medicine — UL1TR002529: Indiana Clinical and Translational Science Institute • Johns Hopkins University — UL1TR003098: Johns Hopkins Institute for Clinical and Translational Research • Loyola Medicine — Loyola University Medical Center • Loyola University Medical Center — UL1TR002389: The Institute for Translational Medicine (ITM) • Maine Medical Center — U54GM115516: Northern New England Clinical & Translational Research (NNE-CTR) Network • Massachusetts General Brigham — UL1TR002541: Harvard Catalyst • Mayo Clinic Rochester — UL1TR002377: Mayo Clinic Center for Clinical and Translational Science (CCaTS) • Medical University of South Carolina — UL1TR001450: South Carolina Clinical & Translational Research Institute (SCTR) • Montefiore Medical Center — UL1TR002556: Institute for Clinical and Translational Research at Einstein and Montefiore • Nemours — U54GM104941: Delaware CTR ACCEL Program • NorthShore University HealthSystem — UL1TR002389: The Institute for Translational Medicine (ITM) • Northwestern University at Chicago — UL1TR001422: Northwestern University Clinical and Translational Science Institute (NUCATS) • OCHIN — INV-018455: Bill and Melinda Gates Foundation grant to Sage Bionetworks • Oregon Health & Science University — UL1TR002369: Oregon Clinical and Translational Research Institute • Penn State Health Milton S. Hershey Medical Center — UL1TR002014: Penn State Clinical and Translational Science Institute • Rush University Medical Center — UL1TR002389: The Institute for Translational Medicine (ITM) • Rutgers, The State University of New Jersey — UL1TR003017: New Jersey Alliance for Clinical and Translational Science • Stony Brook University — U24TR002306 • The Ohio State University — UL1TR002733: Center for Clinical and Translational Science • The State University of New York at Buffalo — UL1TR001412: Clinical and Translational Science Institute • The University of Chicago — UL1TR002389: The Institute for Translational Medicine (ITM) • The University of Iowa — UL1TR002537: Institute for Clinical and Translational Science • The University of Miami Leonard M. Miller School of Medicine — UL1TR002736: University of Miami Clinical and Translational Science Institute • The University of Michigan at Ann Arbor — UL1TR002240: Michigan Institute for Clinical and Health Research • The University of Texas Health Science Center at Houston — UL1TR003167: Center for Clinical and Translational Sciences (CCTS) • The University of Texas Medical Branch at Galveston — UL1TR001439: The Institute for Translational Sciences • The University of Utah — UL1TR002538: Uhealth Center for Clinical and Translational Science • Tufts Medical Center — UL1TR002544: Tufts Clinical and Translational Science Institute • Tulane University — UL1TR003096: Center for Clinical and Translational Science • University Medical Center New Orleans — U54GM104940: Louisiana Clinical and Translational Science (LA CaTS) Center • University of Alabama at Birmingham — UL1TR003096: Center for Clinical and Translational Science • University of Arkansas for Medical Sciences — UL1TR003107: UAMS Translational Research Institute • University of Cincinnati — UL1TR001425: Center for Clinical and Translational Science and Training • University of Colorado Denver, Anschutz Medical Campus — UL1TR002535: Colorado Clinical and Translational Sciences Institute • University of Illinois at Chicago — UL1TR002003: UIC Center for Clinical and Translational Science • University of Kansas Medical Center — UL1TR002366: Frontiers: University of Kansas Clinical and Translational Science Institute • University of Kentucky — UL1TR001998: UK Center for Clinical and Translational Science • University of Massachusetts Medical School Worcester — UL1TR001453: The UMass Center for Clinical and Translational Science (UMCCTS) • University of Minnesota — UL1TR002494: Clinical and Translational Science Institute • University of Mississippi Medical Center — U54GM115428: Mississippi Center for Clinical and Translational Research (CCTR) • University of Nebraska Medical Center — U54GM115458: Great Plains IDeA-Clinical & Translational Research • University of North Carolina at Chapel Hill — UL1TR002489: North Carolina Translational and Clinical Science Institute • University of Oklahoma Health Sciences Center — U54GM104938: Oklahoma Clinical and Translational Science Institute (OCTSI) • University of Rochester — UL1TR002001: UR Clinical & Translational Science Institute • University of Southern California — UL1TR001855: The Southern California Clinical and Translational Science Institute (SC CTSI) • University of Vermont — U54GM115516: Northern New England Clinical & Translational Research (NNE-CTR) Network • University of Virginia — UL1TR003015: iTHRIV Integrated Translational health Research Institute of Virginia • University of Washington — UL1TR002319: Institute of Translational Health Sciences • University of Wisconsin-Madison — UL1TR002373: UW Institute for Clinical and Translational Research • Vanderbilt University Medical Center — UL1TR002243: Vanderbilt Institute for Clinical and Translational Research • Virginia Commonwealth University — UL1TR002649: C. Kenneth and Dianne Wright Center for Clinical and Translational Research • Wake Forest University Health Sciences — UL1TR001420: Wake Forest Clinical and Translational Science Institute • Washington University in St. Louis — UL1TR002345: Institute of Clinical and Translational Sciences • Weill Medical College of Cornell University — UL1TR002384: Weill Cornell Medicine Clinical and Translational Science Center • West Virginia University — U54GM104942: West Virginia Clinical and Translational Science Institute (WVCTSI)

Submitted: Icahn School of Medicine at Mount Sinai — UL1TR001433: ConduITS Institute for Translational Sciences • The University of Texas Health Science Center at Tyler — UL1TR003167: Center for Clinical and Translational Sciences (CCTS) • University of California, Davis — UL1TR001860: UCDavis Health Clinical and Translational Science Center • University of California, Irvine — UL1TR001414: The UC Irvine Institute for Clinical and Translational Science (ICTS) • University of California, Los Angeles — UL1TR001881: UCLA Clinical Translational Science Institute • University of California, San Diego — UL1TR001442: Altman Clinical and Translational Research Institute • University of California, San Francisco — UL1TR001872: UCSF Clinical and Translational Science Institute Pending: Arkansas Children’s Hospital — UL1TR003107: UAMS Translational Research Institute • Baylor College of Medicine — None (Voluntary) • Children’s Hospital of Philadelphia • UL1TR001878: Institute for Translational Medicine and Therapeutics • Cincinnati Children’s Hospital Medical Center — UL1TR001425: Center for Clinical and Translational Science and Training • Emory University — UL1TR002378: Georgia Clinical and Translational Science Alliance • HonorHealth — None (Voluntary) • Loyola University Chicago — UL1TR002389: The Institute for Translational Medicine (ITM) • Medical College of Wisconsin — UL1TR001436: Clinical and Translational Science Institute of Southeast Wisconsin • MedStar Health Research Institute — UL1TR001409: The Georgetown-Howard Universities Center for Clinical and Translational Science (GHUCCTS) • MetroHealth — None (Voluntary) • Montana State University — U54GM115371: American Indian/Alaska Native CTR • NYU Langone Medical Center — UL1TR001445: Langone Health’s Clinical and Translational Science Institute • Ochsner Medical Center — U54GM104940: Louisiana Clinical and Translational Science (LA CaTS) Center • Regenstrief Institute — UL1TR002529: Indiana Clinical and Translational Science Institute • Sanford Research — None (Voluntary) • Stanford University — UL1TR003142: Spectrum: The Stanford Center for Clinical and Translational Research and Education • The Rockefeller University — UL1TR001866: Center for Clinical and Translational Science • The Scripps Research Institute — UL1TR002550: Scripps Research Translational Institute • University of Florida — UL1TR001427: UF Clinical and Translational Science Institute University of New Mexico Health Sciences Center — UL1TR001449: University of New Mexico Clinical and Translational Science Center • University of Texas Health Science Center at San Antonio — UL1TR002645: Institute for Integration of Medicine and Science • Yale New Haven Hospital — UL1TR001863: Yale Center for Clinical Investigation

## Authors statement

Authorship was determined using ICMJE recommendations.

ZB, RH, RW, and CB: Generated research question, drafted manuscript, managed project timeline, and coordinated analysis.

YJ, LW, JA, OO, EH, MK, RCP, AB, ML, JC, AH, JB, LC, and RM: Provided oversight on study design and analysis plan, supported analysis, reviewed manuscript, and provided feedback.

## Inclusion and ethics statement

All co-authors and collaborators included in this manuscript have fulfilled the criteria for authorship.

