## Supplemental Materials for "Metformin and Severe Post-COVID-19 Outcomes Among Individuals with Diabetes Mellitus"

Supplemental Material 1. Rationale for secondary analyses.

Prediabetes is a condition of insulin resistance and moderate hyperglycemia with a high risk of escalation to diabetes.^5,14^ PCOS is an exclusion diagnosis of patients who experience 2 of 3 Rotterdam criteria.^5,15,16^ PCOS is the most common endocrine disorder of reproductive-age women, but the FDA has not approved any medications to treat this condition.^5,15,16^ Prediabetes and PCOS patients are vulnerable to severe COVID-19 outcomes, but are understudied regarding susceptibility to post-COVID-19 outcomes, such as mortality or Long COVID.^5^

Supplemental Material 2. Covariates of interest.

Covariate definitions published previously^23^

"COVID_date",

"long_covid_phenotype",

"visits_per_month",

"data_partner_id",

"BMI_max_observed_or_calculated_before_or_day_of_covid",

"TOBACCOSMOKER_before_or_day_of_covid_indicator",

"OBESITY_before_or_day_of_covid_indicator",

"CHRONICLUNGDISEASE_before_or_day_of_covid_indicator",

"HYPERTENSION_before_or_day_of_covid_indicator",

"DEPRESSION_before_or_day_of_covid_indicator",

"SYSTEMICCORTICOSTEROIDS_before_or_day_of_covid_indicator",

"asthma",

"hba1c",

"insulin",

"HEARTFAILURE_before_or_day_of_covid_indicator",

"DEMENTIA_before_or_day_of_covid_indicator",

"arthritis",

"CORONARYARTERYDISEASE_before_or_day_of_covid_indicator",

"MALIGNANTCANCER_before_or_day_of_covid_indicator",

"METASTATICSOLIDTUMORCANCERS_before_or_day_of_covid_indicator",

"MILDLIVERDISEASE_before_or_day_of_covid_indicator",

"MODERATESEVERELIVERDISEASE_before_or_day_of_covid_indicator",

"KIDNEYDISEASE_before_or_day_of_covid_indicator",

"creatinine",

"ace_inhibitors",

"angiotensin",

"statins",

"anticoagulants",

"aspirin",

"visit_after_enrollment",

"PERIPHERALVASCULARDISEASE_before_or_day_of_covid_indicator",

"CEREBROVASCULARDISEASE_before_or_day_of_covid_indicator",

"glomerular_filtration_rate",

"albumin_creatinine_ratio",

"furosemide",

"torsemide",

"months_from_2018",

"pcos",

"metformin",

"death_within_12_months",

"long_covid_in_study_period",

"death_or_long_covid"

Supplemental Material 3. Characteristics of prediabetes patients prescribed metformin or levothyroxine during acute COVID-19.

| **Characteristic** | **Value** | **Metformin Count (Proportion)** | **Comparator Drugs Count (Proportion)** | **Total Count (Proportion)** |
| --- | --- | --- | --- | --- |
| Total |  | 4296 (0.28) | 11286 (0.72) | 15582 (1) |
| Sex | Female | 2967 (0.69) | 8923 (0.79) | 11890 (0.76) |
| Age: mean (SD) |  | 53.28 (12.64) | 63.09 (12.33) | 60.38 (13.17) |
| Ethnicity | White Non-Hispanic | 2302 (0.54) | 8150 (0.72) | 10452 (0.67) |
|  | Black or African American Non-Hispanic | 896 (0.21) | 923 (0.08) | 1819 (0.12) |
|  | Hispanic or Latino Any Race | 587 (0.14) | 1017 (0.09) | 1604 (0.1) |
|  | Unknown | 249 (0.06) | 553 (0.05) | 802 (0.05) |
|  | Other Non-Hispanic | 26 (0.01) | 67 (0.01) | 93 (0.01) |
|  | American Indian Non-Hispanic | 30 (0.01) | 55 (0) | 85 (0.01) |
|  | Asian or Pacific Islander Non-Hispanic | 206 (0.05) | 521 (0.05) | 727 (0.05) |
| BMI: mean (SD) |  | 38.04 (9.32) | 33.78 (8.45) | 34.9 (8.88) |
| Medical Conditions | Tobacco Smoker | 623 (0.15) | 1729 (0.15) | 2352 (0.15) |
|  | Obese | 3267 (0.76) | 7149 (0.63) | 10416 (0.67) |
|  | Chronic Lung Disease | 909 (0.21) | 3143 (0.28) | 4052 (0.26) |
|  | Hypertension | 2226 (0.52) | 6495 (0.58) | 8721 (0.56) |
|  | Systemic Corticosteroids | 2123 (0.49) | 6657 (0.59) | 8780 (0.56) |
|  | Asthma | 744 (0.17) | 2133 (0.19) | 2877 (0.18) |
|  | Heart Failure | 187 (0.04) | 950 (0.08) | 1137 (0.07) |
|  | Dementia | 38 (0.01) | 248 (0.02) | 286 (0.02) |
|  | Arthritis | 66 (0.02) | 413 (0.04) | 479 (0.03) |
|  | Coronary Artery Disease | 321 (0.07) | 1559 (0.14) | 1880 (0.12) |
|  | Cancer | 348 (0.08) | 2151 (0.19) | 2499 (0.16) |
|  | Liver Disease | <20 (<20) | N/A (N/A) | N/A (N/A) |
|  | Chronic Kidney Disease | 134 (0.03) | 610 (0.05) | 744 (0.05) |
|  | Peripheral Vascular Disease | 87 (0.02) | 448 (0.04) | 535 (0.03) |
|  | Cerebrovascular Disease | 134 (0.03) | 948 (0.08) | 1082 (0.07) |
|  | Depression | 1189 (0.28) | 3488 (0.31) | 4677 (0.3) |
| Drugs | Insulin | 249 (0.06) | 789 (0.07) | 1038 (0.07) |
|  | Ace Inhibitors | 236 (0.05) | 373 (0.03) | 609 (0.04) |
|  | Angiotensin Receptor Blockers | 168 (0.04) | 361 (0.03) | 529 (0.03) |
|  | Statins | 283 (0.07) | 697 (0.06) | 980 (0.06) |
|  | Anticoagulants | 214 (0.05) | 769 (0.07) | 983 (0.06) |
|  | Aspirin | 250 (0.06) | 723 (0.06) | 973 (0.06) |
|  | Torsemide | 22 (0.01) | 70 (0.01) | 92 (0.01) |
|  | Furosemide | 317 (0.07) | 1161 (0.1) | 1478 (0.09) |
| Measurements | Pre-Prescription Hba1c: mean (SD) | 5.95 (0.51) | 5.76 (0.33) | 5.81 (0.39) |
|  | Serum Creatinine: mean (SD) | 0.79 (0.27) | 0.83 (0.25) | 0.82 (0.25) |
|  | Albumin/Creatinine Ratio: mean (SD) | 7.77 (25.96) | 7.52 (38.35) | 7.6 (34.61) |
|  | Glomerular Filtration Rate: mean (SD) | 76.01 (26.09) | 71.8 (20.91) | 72.79 (22.31) |
| Medical Utilization | Visits per Month: mean (SD) | 2.02 (2.33) | 2.63 (2.77) | 2.47 (2.67) |
| Outcomes | Long COVID Positivity | 56 (0.01) | 208 (0.02) | 264 (0.02) |

We evaluated EHR from a sample of 15,582 individuals with prediabetes and comorbid COVID-19. Of these patients, 4,269 were prescribed metformin before and during acute COVID-19 (mean age 53 years), while 11,286 were prescribed levothyroxine (mean age 63 years).

Supplemental Material 4. Characteristics of PCOS patients prescribed metformin or levothyroxine during acute COVID-19.

| Characteristic | Value | Metformin Count (Proportion) | Comparator Drugs Count (Proportion) | Total Count (Proportion) |
| --- | --- | --- | --- | --- |
| Total |  | 1135 (0.64) | 651 (0.37) | 1786 (1) |
| Age: mean (SD) |  | 38.43 (6.91) | 40.48 (8.16) | 39.18 (7.46) |
| Ethnicity | White Non-Hispanic | 736 (0.65) | 479 (0.74) | 1215 (0.68) |
|  | Black or African American Non-Hispanic | 154 (0.14) | 31 (0.05) | 185 (0.1) |
|  | Hispanic or Latino Any Race | 133 (0.12) | 53 (0.08) | 186 (0.1) |
|  | Unknown | 56 (0.05) | 44 (0.07) | 100 (0.06) |
|  | Other Non-Hispanic | <20 (<20) | <20 (<20) | <20 (<20) |
|  | American Indian Non-Hispanic | <20 (<20) | <20 (<20) | <20 (<20) |
|  | Asian or Pacific Islander Non-Hispanic | 44 (0.04) | 40 (0.06) | 84 (0.05) |
| BMI: mean (SD) |  | 40.64 (9.88) | 38.59 (10.32) | 39.87 (10.1) |
| Medical Conditions | Tobacco Smoker | 136 (0.12) | 76 (0.12) | 212 (0.12) |
|  | Obese | 920 (0.81) | 485 (0.75) | 1405 (0.79) |
|  | Chronic Lung Disease | 244 (0.21) | 176 (0.27) | 420 (0.24) |
|  | Hypertension | 287 (0.25) | 177 (0.27) | 464 (0.26) |
|  | Systemic Corticosteroids | 547 (0.48) | 397 (0.61) | 944 (0.53) |
|  | Asthma | 250 (0.22) | 172 (0.26) | 422 (0.24) |
|  | Heart Failure | <20 (<20) | <20 (<20) | 24 (0.01) |
|  | Dementia | <20 (<20) | <20 (<20) | <20 (<20) |
|  | Arthritis | <20 (<20) | <20 (<20) | 32 (0.02) |
|  | Coronary Artery Disease | <20 (<20) | <20 (<20) | 26 (0.01) |
|  | Cancer | 37 (0.03) | 63 (0.1) | 100 (0.06) |
|  | Liver Disease | <20 (<20) | <20 (<20) | <20 (<20) |
|  | Chronic Kidney Disease | 20 (0.02) | 25 (0.04) | 45 (0.03) |
|  | Peripheral Vascular Disease | <20 (<20) | <20 (<20) | <20 (<20) |
|  | Cerebrovascular Disease | <20 (<20) | <20 (<20) | <20 (<20) |
|  | Depression | 397 (0.35) | 234 (0.36) | 631 (0.35) |
| Drugs | Insulin | 40 (0.04) | 33 (0.05) | 73 (0.04) |
|  | Ace Inhibitors | N/A (N/A) | <20 (<20) | N/A (N/A) |
|  | Angiotensin Receptor Blockers | <20 (<20) | <20 (<20) | <20 (<20) |
|  | Statins | <20 (<20) | <20 (<20) | <20 (<20) |
|  | Anticoagulants | 35 (0.03) | 22 (0.03) | 57 (0.03) |
|  | Torsemide | 0 (0) | 0 (0) | 0 (0) |
|  | Furosemide | 29 (0.03) | 27 (0.04) | 56 (0.03) |
| Measurements | Pre-Prescription Hba1c: mean (SD) | 5.48 (0.48) | 5.31 (0.4) | 5.42 (0.46) |
|  | Serum Creatinine: mean (SD) | 0.69 (0.25) | 0.75 (0.21) | 0.71 (0.24) |
|  | Albumin/Creatinine Ratio: mean (SD) | 30.13 (62.52) | 2.5 (1.9) | 26.94 (59.47) |
|  | Glomerular Filtration Rate: mean (SD) | 79.91 (27.1) | 79.05 (20.57) | 79.55 (24.54) |
| Medical Utilization | Visits per Month: mean (SD) | 2.03 (2.7) | 2.73 (3.89) | 2.29 (3.2) |
| Outcomes | Long COVID Positivity | <20 (<20) | <20 (<20) | 30 (0.02) |

We evaluated a sample of 1,768 individuals with PCOS and comorbid COVID-19. Of these individuals, 1,135 were prescribed metformin (mean age 38 years) before and during acute COVID-19, while 651 were prescribed levothyroxine (mean age 40 years).

Supplemental Material 5. Relationships between metformin vs. levothyroxine during acute COVID-19 and subsequent 12-month cumulative incidence of Long COVID or death, among PCOS and prediabetes patients.

Supplemental Material 5a. Adjusted relationships between metformin vs. levothyroxine during acute COVID-19 and subsequent 12-month cumulative incidence of Long COVID or death, among individuals with PCOS or prediabetes.

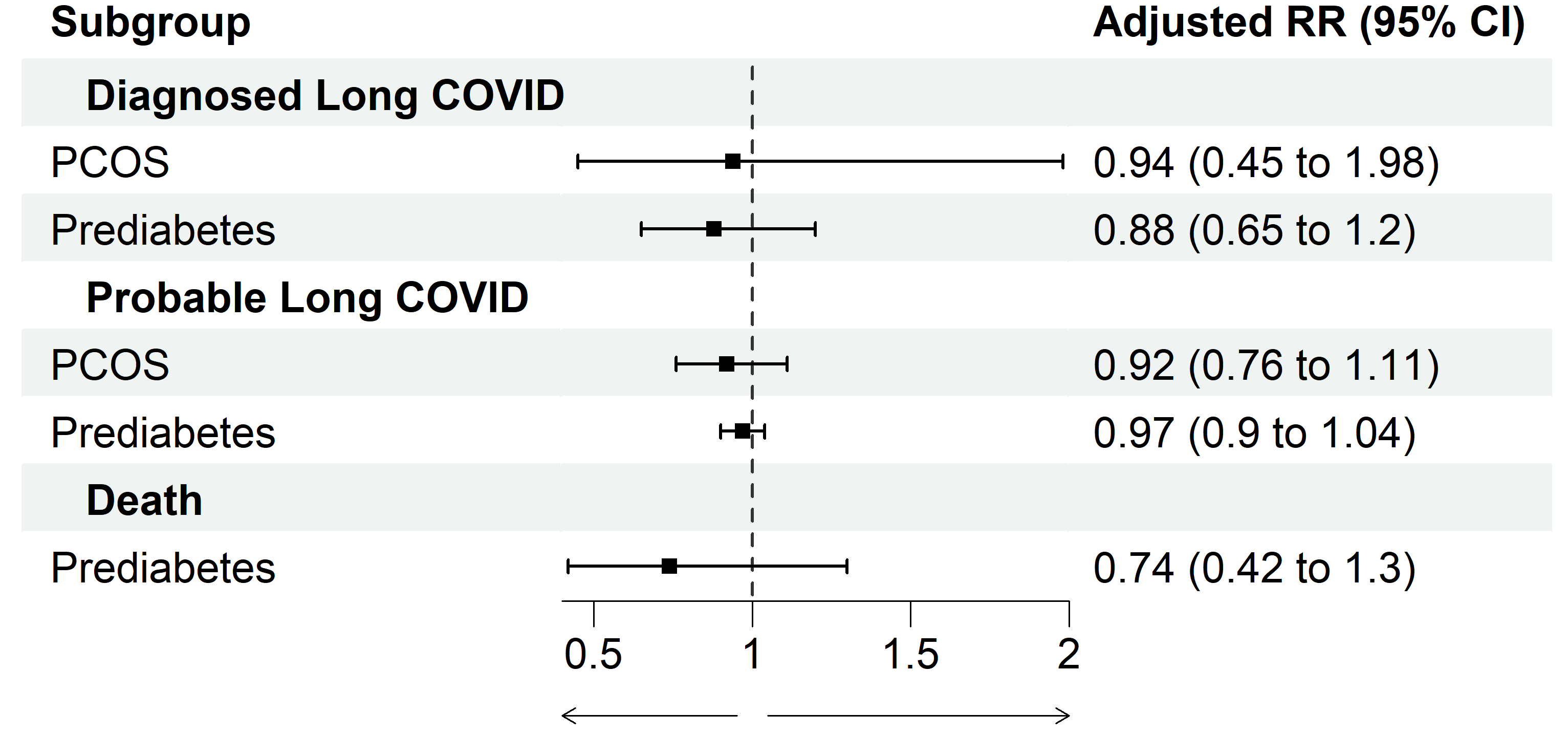

Supplemental Material 5b. Unadjusted relationships between metformin vs. levothyroxine during acute COVID-19 and subsequent 12-month cumulative incidence of Long COVID or death, among individuals with PCOS or prediabetes.

|  | Sample size Metformin | Sample size Levothyroxine | Metformin Outcomes | Levothyroxine Outcomes | Metformin Unadjusted risk | Levothyroxine Unadjusted Risk | Unadjusted RR (95% CI) |
| --- | --- | --- | --- | --- | --- | --- | --- |
| PCOS/Long COVID | 1135 | 651 | 18 | 12 | 0.02 | 0.02 | 0.86 (0.42 to 1.77) |
| Prediabetes/Long COVID | 4296 | 11286 | 56 | 208 | 0.01 | 0.02 | 0.71 (0.53 to 0.95) |
| PCOS/Long COVID Phenotype | 1135 | 651 | 189 | 138 | 0.17 | 0.21 | 0.79 (0.64 to 0.96) |
| Prediabetes/Long COVID Phenotype | 4296 | 11286 | 734 | 2503 | 0.17 | 0.22 | 0.77 (0.72 to 0.83) |
| PCOS/Death | 1135 | 651 | 0 | 0 | 0.00 | 0.00 | N/A (N/A to N/A) |
| Prediabetes/Death | 4296 | 11286 | 25 | 233 | 0.01 | 0.02 | 0.28 (0.19 to 0.43) |

Among individuals with PCOS and comorbid COVID-19, we found that prescription of metformin, compared to levothyroxine, was not significantly associated with the one-year cumulative incidence of LC-Dx (aRR 0.95, 95% CI 0.45, 1.98) or LC-P (aRR 0.92, 95% CI 0.77, 1.11) (mortality confidence interval did not converge).

Among individuals with prediabetes and comorbid COVID-19, prescription of metformin compared to levothyroxine was not significantly associated with the one-year cumulative incidence of LC-Dx (aRR 0.88, 95% CI 0.65, 1.20), LC-P (aRR 0.97, 95% CI 0.90, 1.04), or mortality (aRR 0.74, 95% CI 0.42, 1.30).

Supplemental Material 6. Discussion of secondary cohort findings.

We did not find a significant relationship between the prescription of metformin, compared to levothyroxine, and long-term consequences of COVID-19 among patients with prediabetes or PCOS. Given the small sample size of individuals with prediabetes and PCOS in our analyses, it is unclear whether the protective effects of metformin against the negative long-term consequences of COVID-19 are limited to T2DM patients.
